# Prevalence of anticipatory nausea and vomiting in children undergoing cytotoxic chemotherapy for malignant disease - the SiCK2 (Sickness prior to Chemotherapy in Kids) study

**DOI:** 10.1101/2020.06.09.20126284

**Authors:** Bob Phillips, Patric ffrench-Devitt, Lucy Wellings

## Abstract

**Introduction:** Anticipatory nausea and vomiting (ANV) is thought to be a conditioned response to nausea and vomiting experienced in previous chemotherapy cycles, and has a significant negative impact on quality of life for children having treatment for cancer. The prevalence of this aversive experience with current antiemetics remains uncertain.

**Methods:** Self-report questionnaires completed by patients and parents across seven sites in the UK. Nausea and vomiting symptoms 24 hours prior to commencing chemotherapy were assessed with the Pediatric Nausea Assessment Tool (PeNAT). Data were also collected on the patients’ age, sex, oncological diagnosis, and previous experience of chemotherapy induced nausea and vomiting. Correlation between demographic data, chemotherapy information and prior reported experience of chemotherapy related nausea and vomiting was undertaken using multiple ordinal regression.

**Results:** 191 episodes of anticipatory nausea and vomiting status were returned. 34% of patients described severe or very severe anticipatory nausea and/or vomiting. The severity of anticipatory nausea and vomiting was predicted two factors related to prior chemotherapy: control of anticipatory (OR 0.23 95%CI 0.09 to 0.53) and acute/delayed (OR 0.37, 95% CI 0.16 to 0.83) nausea and vomiting, and one current factor, the administration of antiemetic medication prior to arrival at the hospital (OR 3.0, 95%CI 1.3 to 6.8).

**Conclusions:** This study re-enforces, disappointingly, the continued high prevalence of anticipatory nausea and vomiting in children about to receive chemotherapy. There is clearly a need to improve interventions for this rarely discussed aversive experience of childhood cancer. Its high prevalence suggests trials of interventions should be possible to power effectively, and develop interventions that are both acceptable and deliverable.

## 1. INTRODUCTION

Patients undergoing cytotoxic chemotherapy as part of cancer treatment often develop numerous side effects, including nausea and vomiting. Nausea and vomiting symptoms have a significant negative impact on quality of life for children having treatment for cancer^1^. The phenomenon of “anticipatory” nausea and vomiting (i.e. that which develops prior to the administration of chemotherapeutic agents) has been described. Despite availability of modern anti-emetics, it is estimated to affect around 10% of adults receiving cancer treatment, with nausea more common than vomiting^2^. Previous studies into the prevalence of anticipatory nausea and vomiting in children have been problematic, as a lack of validated symptom scores and questionnaires have resulted in many taking parental reports of their child’s symptoms as a proxy measure of symptom occurrence and severity^3^. An international guideline for the management of anticipatory nausea and vomiting in children specifically highlights the lack of appropriate literature surrounding the prevalence of this phenomenon in this population^4^.

Anticipatory nausea and vomiting is thought to be a conditioned response to nausea and vomiting experienced in previous chemotherapy cycles ^5^. This theory proposes children who have previously experienced nausea and vomiting during chemotherapy will re-experience these symptoms when they know they will soon be exposed to another cycle of chemotherapy, even before the infusion is commenced or tablets have been swallowed. Following from this, it’s predicted improved control of chemotherapy induced nausea and vomiting (CINV) will reduce prevalence of anticipatory nausea and vomiting. CINV is often inadequately controlled in children and while has been postulated that children experience higher levels of anticipatory nausea and vomiting than adults^6^ a previous small scale study suggested that levels are similar to those found in adults7. Given the small size and other study limitations it is difficult to know whether this is the true prevalence and as such it is difficult to know how much emphasis should be put on its prevention and management.

This project aimed to determine the prevalence of anticipatory nausea and vomiting amongst children and young people undergoing cytotoxic chemotherapy in tertiary oncology centres in the UK. Given past data, we hypothesised the rate of nausea and vomiting may be influenced by gender, age, experience of and emetogenicity of previous chemotherapy.

## 2. METHODS

A questionnaire-based study, delivered directly to parents and patients over a six week period in 2018 was undertaken across seven sites in the UK. The postcard-sized questionnaire assessed nausea and vomiting symptoms 24 hours prior to commencing a new chemotherapy cycle for with implied consent by return of the card. The Pediatric Nausea Assessment Tool (PeNAT) was be used to assess symptoms of nausea in children aged 4-18, with caregiver reporting in children under 4 years old. Data were also collected on the patients’ age, sex, oncological diagnosis, and previous experience of chemotherapy induced nausea and vomiting.

Patients were eligible for inclusion if they were aged 18 or under and were receiving chemotherapy for a cancer or cancer-like diagnosis (for example, a low grade glioma brain tumour of haemophagocytic lymphohistiocytosis), and agreed to complete the questionnaire.

The analysis examined the overall prevalence of anticipatory nausea and vomiting, and correlating its’ severity with demographic data, chemotherapy information and prior reported experience of CINV using multiple ordinal regression.

This research was supported with funding from Candlelighters Trust, and approved by the UK National health research ethics board (IRAS ID 232500).

## 3. RESULTS

Data from 219 episodes of chemotherapy were returned, of which 191 reported their anticipatory nausea and vomiting status. The median age of the patients was 7 years (range 0.3 to 18), and 40% were female. The most common diagnosis was acute lymphoblastic leukaemia (ALL) of which 94% were in the maintenance phase – see Table 1. 93% of patients had received chemotherapy prior to the reported cycle.

**Table 1:**
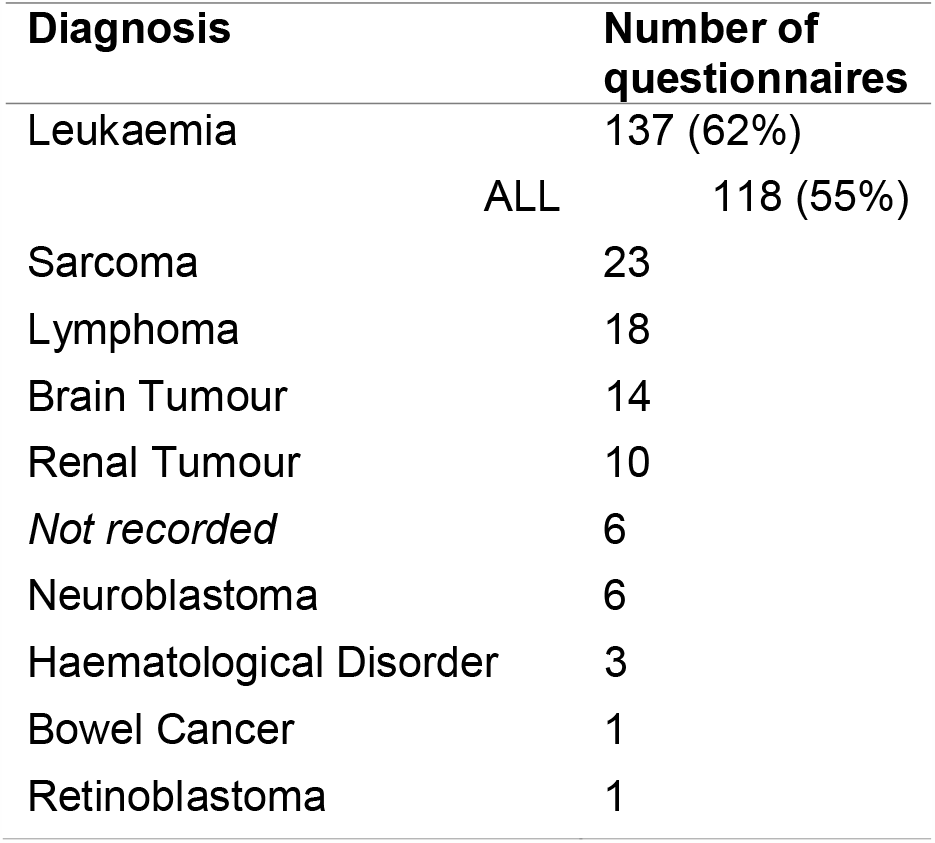
Diagnosis of returned questionnaires

The majority of patients did not report anticipatory nausea and vomiting (median PeNAT score 1, range 1-4), but importantly 17% (33/191) had severe (3) or very severe (4) nausea before chemotherapy and 17% (34/191) had vomited at least once. (See Figure 1.)

**Figure 1:**
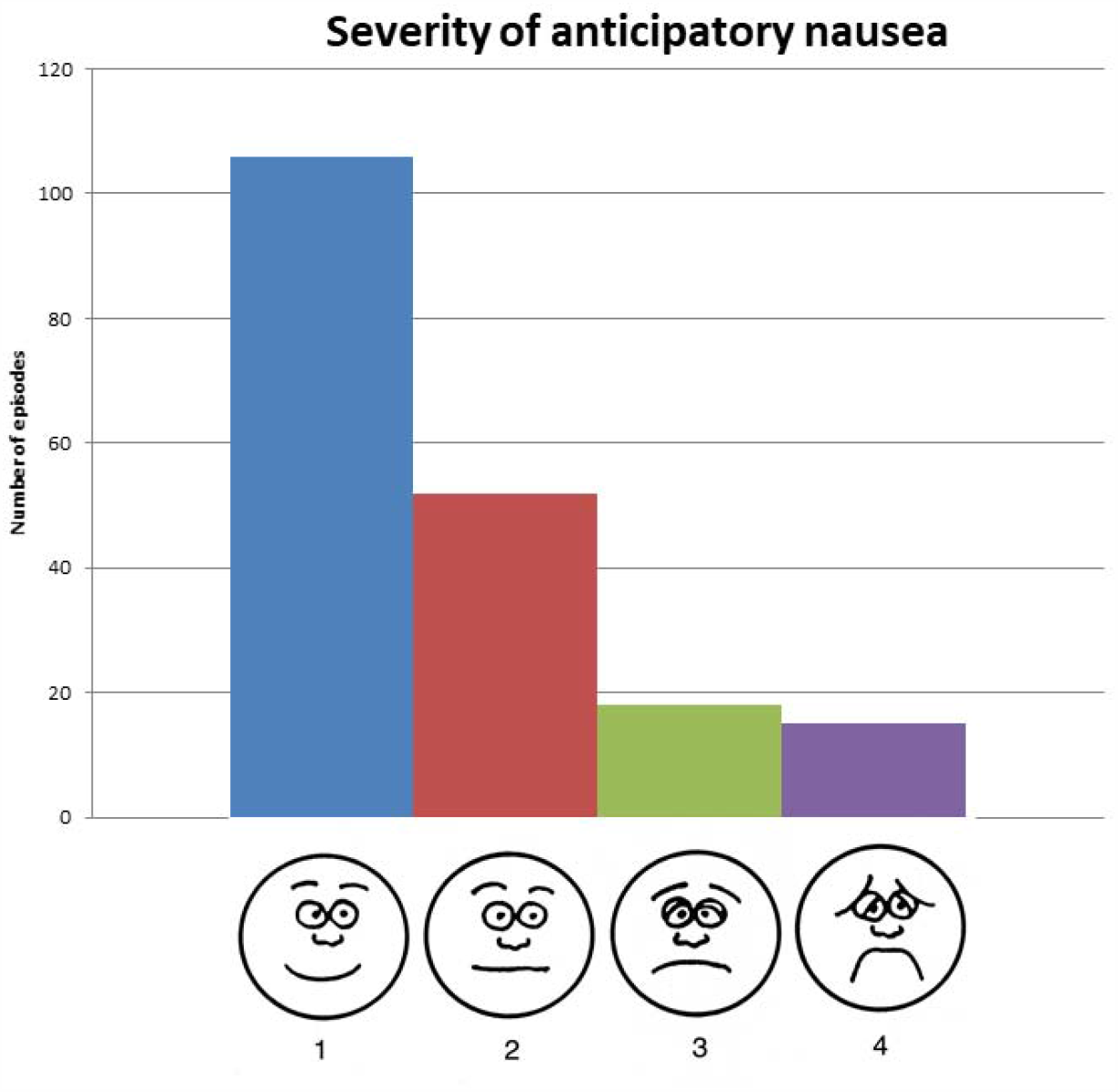
Anticipatory nausea scores

On multivariable analysis, the severity of anticipatory nausea and vomiting being experienced was beneficially predicted by control of nausea and vomiting before (OR 0.23 95%CI 0.09 to 0.53) or during (OR 0.37, 95% CI 0.16 to 0.83) prior chemotherapy, and adversely if the patient had been given antiemetic medication prior to arrival at the hospital (OR 3.0, 95%CI 1.3 to 6.8). Sex, age, their interaction, and type of diagnosis were not found to be independently predictive.

## 4. DISCUSSION

This simple study updates and re-enforces disappointingly the continued high prevalence of anticipatory nausea and vomiting in children about to receive chemotherapy, despite the improvement of antiemetics, and the high proportion of those receiving minimally emetogenic chemotherapy (ALL maintenance). Our study uses a well validated measure to assess this problem, and emphasises the previously assessed factors of poor control experience in previous cycles. It also demonstrates that beyond this, those families which pre-medicate their children have a higher reported severity of anticipatory nausea and vomiting.

The study does not cover all UK centres, and despite its large number of respondents, does not have adequate power to truly separate the emetogenicity of the chemotherapy about to be undertaken and knowledge of this with the anticipated experience.

Our study emphasises again the need to rigorously adhere to antiemetic prophylaxis strategies during chemotherapy. Undertaking this would improve acute control^8^ as well as anticipatory, and potentially other side effects such as weight loss and febrile episodes^9^.

There is clearly a need to improve interventions for this rarely discussed aversive experience of childhood cancer. Current guidance of secondary prevention is based on only small studies with limited interventions, suggesting desensitising and hypnosis techniques and low doses of benzodiazepine.^4^ The high prevalence of anticipatory nausea and vomiting suggests trials of interventions should be possible to power effectively, and develop interventions that are both acceptable and deliverable.^10^

## Data Availability

Data available for collaborative projects on request

